# Intracardiac Thrombus in COVID-19 Inpatients: A Nationwide Study of Incidence, Predictors and Outcomes

**DOI:** 10.1101/2023.07.05.23292278

**Authors:** Ankit Agrawal, Suryansh Bajaj, Umesh Bhagat, Sanya Chandna, Aro Daniela Arockiam, Nicholas Chan, Elio Haroun, Rahul Gupta, Osamah Badwan, Shashank Shekhar, Shivabalan Kathavarayan Ramu, Divya Nayar, Wael Jaber, Brian P Griffin, Tom Kai Ming Wang

## Abstract

**Background:** COronaVIrus Disease 2019 (COVID-19) has been observed to be associated with a hypercoagulable state. Intracardiac thrombosis is a serious complication but has seldom been evaluated in COVID-19 patients. We assessed the incidence, associated factors, and outcomes of COVID-19 patients with intracardiac thrombosis.

**Methods:** COVID-19 inpatients during 2020 were retrospectively identified from the national inpatient sample (NIS) database, and data retrieved regarding clinical characteristics, intracardiac thrombosis, and adverse outcomes. Multivariable logistic regression was performed to identify the clinical factors associated with intracardiac thrombosis and in-hospital mortality and morbidities.

**Results:** A total of 1,683,785 COVID-19 inpatients were identified in 2020 from NIS, with a mean age of 63.8 ± 1.6 years, and 32.2% females. Intracardiac thrombosis was present in 0.001% (1,830) patients. Overall, in-hospital outcomes include all-cause mortality 13.2% (222,695/1,683,785), cardiovascular mortality 3.5%, cardiac arrest 2.6%, acute coronary syndrome (ACS) 4.4%, heart failure 16.1%, stroke 1.3% and acute kidney injury (AKI) 28.3%. The main factors associated with intracardiac thrombosis were a history of congestive heart failure and coagulopathy. Intracardiac thrombosis was independently associated with a higher risk of in-hospital all-cause mortality (OR: 3.32, 95% CI: 2.42-4.54, p<0.001), cardiovascular mortality (OR: 2.95, 95% CI: 1.96-4.44, p<0.001), cardiac arrest (OR: 2.04, 95% CI: 1.22-3.43, p=0.006), ACS (OR: 1.62, 95% CI: 1.17-2.22, p=0.003), stroke (OR: 3.10, 95% CI: 2.11-4.56, p<0.001), and AKI (OR: 2.13 95% CI: 1.68-2.69, p<0.001), but not incident heart failure (p=0.27).

**Conclusion:** Although intracardiac thrombosis is rare in COVID-19 inpatients, its presence was independently associated with higher risks of in-hospital mortality and most morbidities. Prompt investigations and treatments for intracardiac thrombosis are warranted when there is a high index of suspicion and a confirmed diagnosis respectively.

## INTRODUCTION

COVID-19 was declared as a pandemic by the World Health Organization (WHO) in March 2020.^1^ While the culprit virus primarily infects the respiratory tract, there is growing evidence of the multisystem involvement of the illness. Recent studies have shown that the recent strains of coronavirus promote a hypercoagulable state via abnormally elevated levels of procoagulant cytokines in circulation and virus-mediated endothelial inflammation.^2, 3^ The thrombotic sequala varies in terms of location and clinical presentation but is often associated with adverse clinical outcomes. There are a few case reports and series of intracardiac thrombosis in COVID-19 patients, many of whom with no prior history of cardiovascular disease or risk factors for coagulopathy.^4–8^ While some studies have estimated the incidence and outcomes of arterial and venous thromboembolism in COVID-19 patients, data are lacking regarding intracardiac thrombosis in COVID-19 patients. To the best of our knowledge, this is the first large-scale nationwide study to assess the incidence of and factors associated with intracardiac thrombosis in COVID-19 patients and evaluate its association with end-organ damage and patient mortality.

## METHODS

### Study population

The National Inpatient Sample (NIS) database 2020, managed by the Agency for Healthcare Research and Quality via Healthcare Cost and Utilization Project (HCUP),^9^ was used for the collection of patient data. The NIS database is a collection of over 7 million inpatient data reported annually from all the participating states. Publicly available and anonymous data from the NIS website without any patient identifying features were used for our study, which precludes the requirement of Institutional Review Board approval and informed consent.

### Study characteristics and outcomes

International Classification of Diseases, Tenth Revision, Clinical Modification (ICD-10-CM), claims codes were used to identify the patient cohort. Firstly, patients hospitalized for COVID-19 were identified using ICD-10-CM code U07.1. Amongst this patient cohort, ICD-10-CM code I51.3 was used to identify patients with concomitant intracardiac thrombosis diagnosed during hospitalization. Patients aged<18 years were excluded from our study.

The following NIS variables were used for data collection: age, sex, race, hospital type, and hospital region. Conditions such as congestive heart failure, valvular disease, pulmonary circulation disorders, peripheral vascular disorder, uncomplicated and complicated hypertension, paralysis, other neurological disorder, chronic obstructive pulmonary disorder, uncomplicated and complicated diabetes, renal failure, liver disease, lymphoma, metastatic cancer, solid tumor, acquired immune deficiency syndrome/human immunodeficiency virus, rheumatoid arthritis/collagen vascular disorder, blood loss anemia, deficiency anemia, and alcohol abuse were part of the Elixhauser comorbidity index and were utilized directly in the analysis. ICD-10-CM codes were used to identify the remaining variables. Asymptomatic COVID-19 was defined as patients with the diagnosis of COVID-19 without any related symptoms. Symptomatic COVID-19 was defined as patients with COVID-19 and acute bronchitis, lower respiratory tract infection, pneumonia, and acute respiratory distress syndrome (ARDS). Severe COVID-19 was defined as patients with COVID-19 and related severe sepsis and/or septic shock, intubation and assisted ventilation, non-invasive ventilation, use of extracorporeal membrane oxygenation, cardiac arrest, acute respiratory failure, need for hemodialysis and packed red blood cell transfusion.

The primary endpoint was intracardiac thrombosis. The secondary endpoints studied were in-hospital all-cause and cardiovascular mortality and in-hospital complications, including acute kidney injury (AKI), stroke, cardiac arrest, acute coronary syndrome (ACS), and heart failure.

### Statistical Analyses

Means with standard deviations (or medians with interquartile ranges) and number of individuals with proportions were calculated for continuous and categorical variables, respectively. The characteristics between those with and without intracardiac thrombosis were compared using student t-test (or Wilcoxon Rank-Sum test for non-normal distribution, with normality assessed by Shapiro-Wilk test) for continuous variables and chi-square test (or Fisher exact test) for categorical variables. Multivariate logistic regression analysis was performed to identify the risk factors for organ damage and patient mortality in the overall cohort, and with intracardiac thrombosis. All the covariates utilized are risk factors based on clinical significance from established literature. A p-value of <0.05 was considered statistically significant. The analysis was conducted using StataCorp. 2021. Stata Statistical Software: Release 17. StataCorp LLC (College station, TX, USA). Stata’s every command and appropriate weights were used in all estimations.

## RESULTS

### Baseline characteristics

A total of 1,683,875 patients who tested positive for COVID-19 in the year 2020 were identified, out of which, 1,830 patients were found to have intracardiac thrombosis. The incidence was 0.001% with a mean age of 63.8 ±1.6 years and 32.2% females. Demographic and clinical characteristics of COVID-19 patients with and without intracardiac thrombosis are compared and described in Table 1.Compared to COVID patients without intracardiac thrombosis, those with thrombosis were more likely to be men (67.8% vs.52%, p<0.001), African Americans (31.5% vs 19%, p<0.001), have a longer median length of stay in the hospital (8 (4-14) vs.5 (3-10) days), be asymptomatic (34.5% vs. 24.8%. p<0.001), and admitted to large (58% vs. 46.8%, p<0.001) urban teaching hospitals (84.4% vs. 71.8%, p<0.001). The in-hospital all-cause mortality and cardiovascular mortality in the overall cohort were 13.2% (222,695/1,683,785) and 3.5% (59530/1,683,785) respectively; the in-hospital morbidity rate was 35.8% (602,795/1,683,785). The incidence of in-hospital mortality and complications in COVID-19 patients without intracardiac thrombosis were significantly lower than COVID-19 patients with intracardiac thrombosis (Table 2).

**Table 1:**
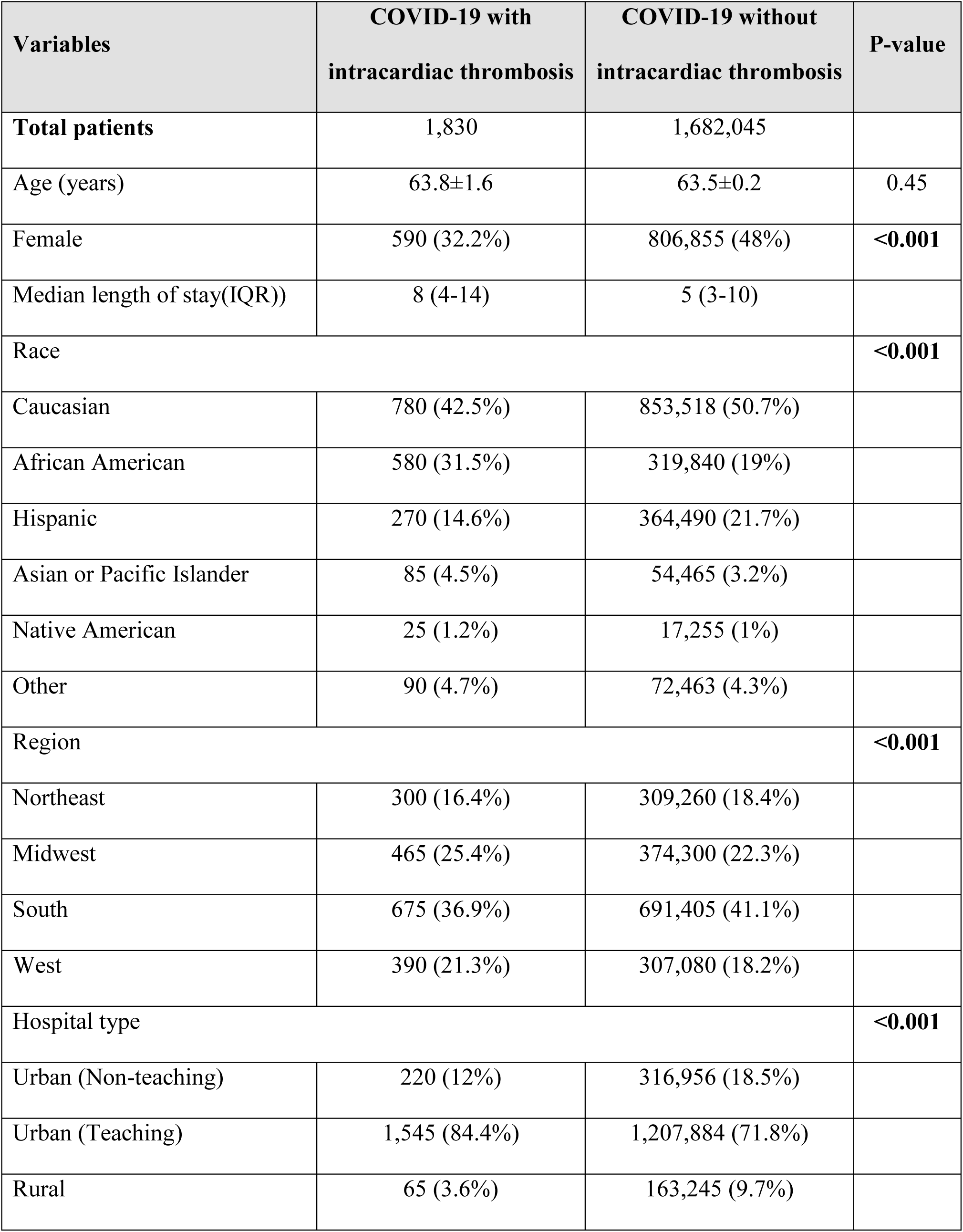
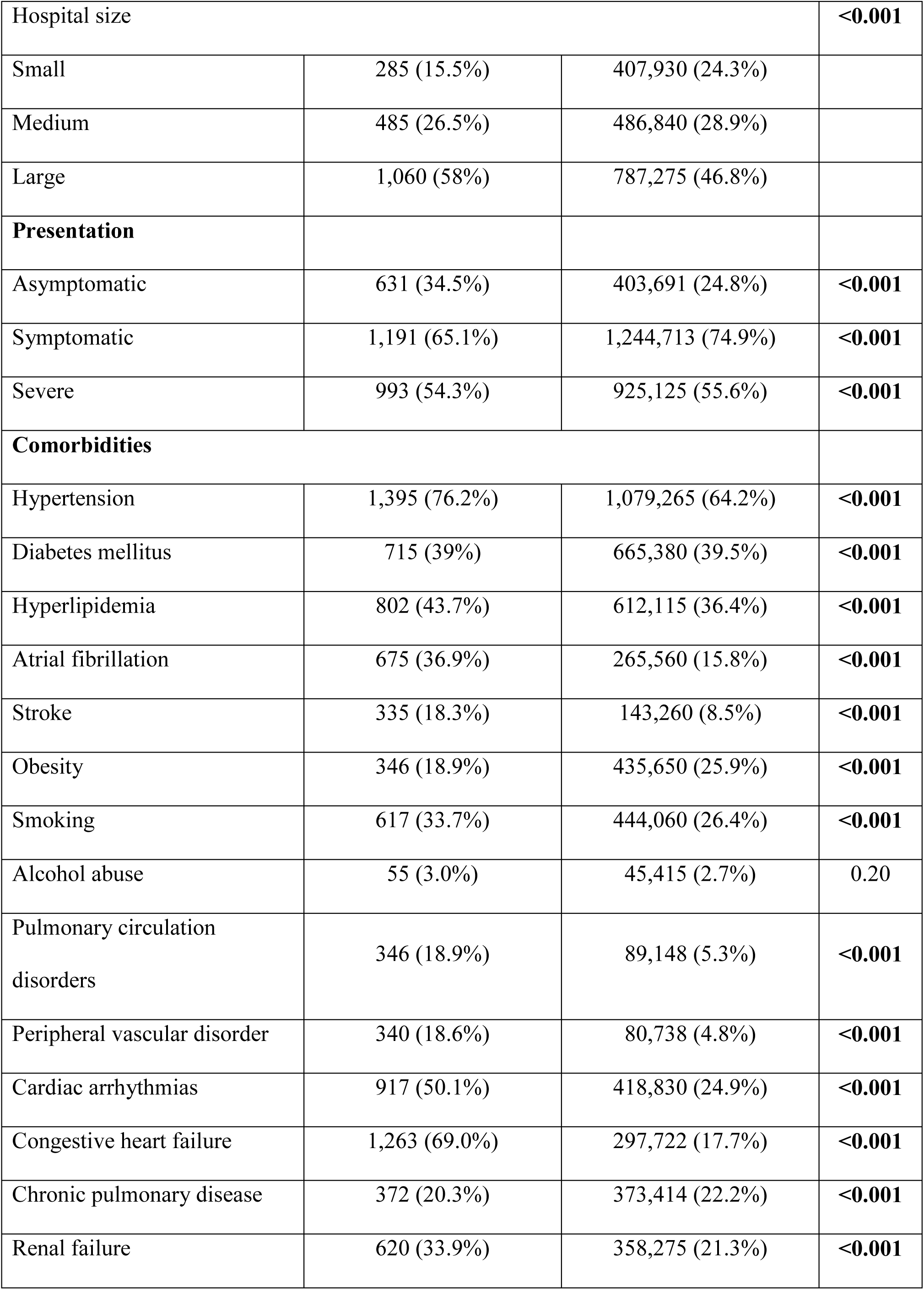
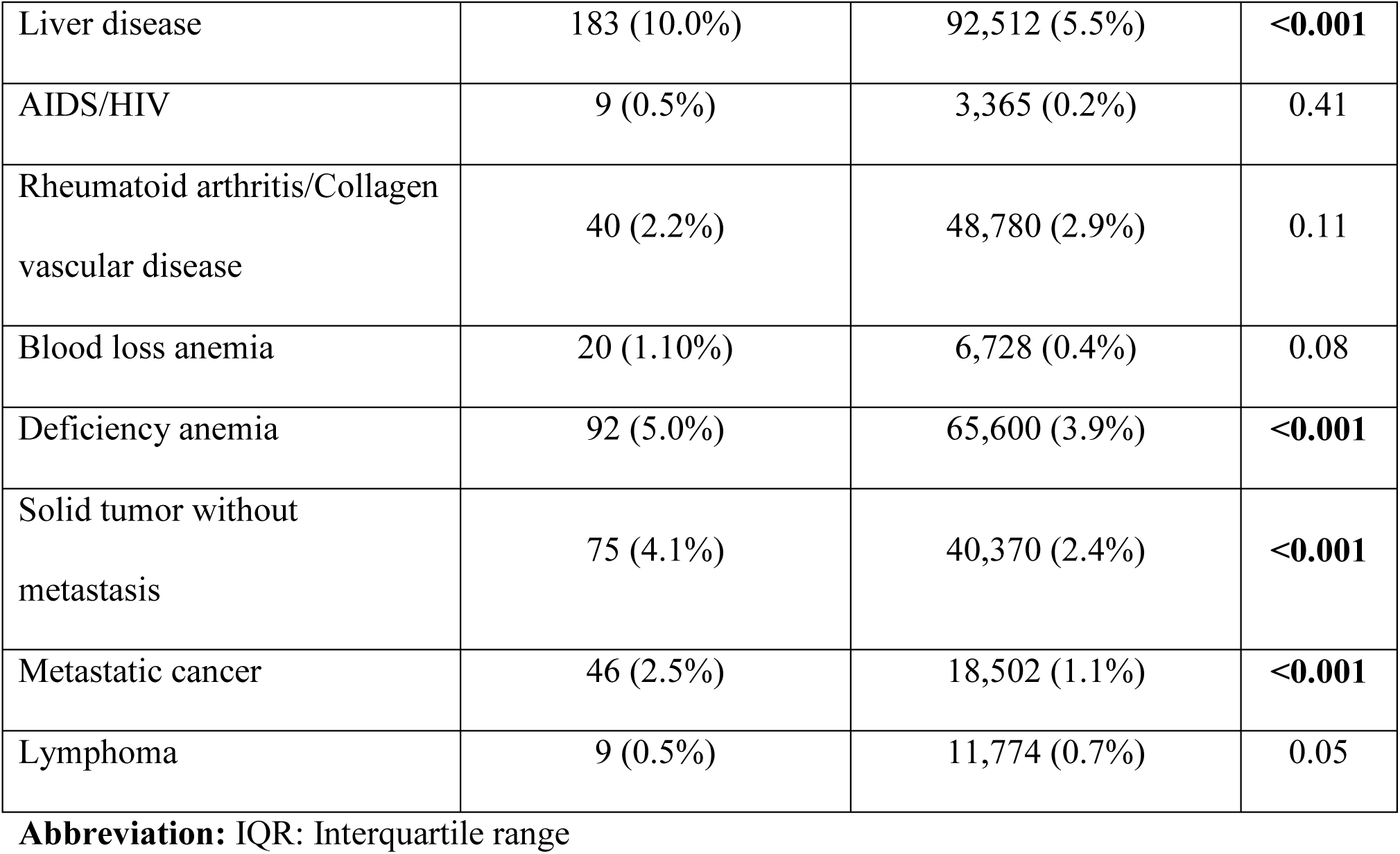
Baseline characteristics of COVID-19 patients with and without intracardiac thrombosis.

| Variables | COVID-19 with intracardiac thrombosis | COVID-19 without intracardiac thrombosis | P-value |
| --- | --- | --- | --- |
| Total patients | 1,830 | 1,682,045 |  |
| Age (years) | 63.8±1.6 | 63.5±0.2 | 0.45 |
| Female | 590 (32.2%) | 806,855 (48%) | <0.001 |
| Median length of stay(IQR)) | 8 (4-14) | 5 (3-10) |  |
| Race |  |  | <0.001 |
| Caucasian | 780 (42.5%) | 853,518 (50.7%) |  |
| African American | 580 (31.5%) | 319,840 (19%) |  |
| Hispanic | 270 (14.6%) | 364,490 (21.7%) |  |
| Asian or Pacific Islander | 85 (4.5%) | 54,465 (3.2%) |  |
| Native American | 25 (1.2%) | 17,255 (1%) |  |
| Other | 90 (4.7%) | 72,463 (4.3%) |  |
| Region |  |  | <0.001 |
| Northeast | 300 (16.4%) | 309,260 (18.4%) |  |
| Midwest | 465 (25.4%) | 374,300 (22.3%) |  |
| South | 675 (36.9%) | 691,405 (41.1%) |  |
| West | 390 (21.3%) | 307,080 (18.2%) |  |
| Hospital type |  |  | <0.001 |
| Urban (Non-teaching) | 220 (12%) | 316,956 (18.5%) |  |
| Urban (Teaching) | 1,545 (84.4%) | 1,207,884 (71.8%) |  |
| Rural | 65 (3.6%) | 163,245 (9.7%) |  |
| Hospital size |  |  | **<0.001** |
| Small | 285 (15.5%) | 407,930 (24.3%) |  |
| Medium | 485 (26.5%) | 486,840 (28.9%) |  |
| Large | 1,060 (58%) | 787,275 (46.8%) |  |
| **Presentation** |  |  |  |
| Asymptomatic | 631 (34.5%) | 403,691 (24.8%) | **<0.001** |
| Symptomatic | 1,191 (65.1%) | 1,244,713 (74.9%) | **<0.001** |
| Severe | 993 (54.3%) | 925,125 (55.6%) | **<0.001** |
| **Comorbidities** |  |  |  |
| Hypertension | 1,395 (76.2%) | 1,079,265 (64.2%) | **<0.001** |
| Diabetes mellitus | 715 (39%) | 665,380 (39.5%) | **<0.001** |
| Hyperlipidemia | 802 (43.7%) | 612,115 (36.4%) | **<0.001** |
| Atrial fibrillation | 675 (36.9%) | 265,560 (15.8%) | **<0.001** |
| Stroke | 335 (18.3%) | 143,260 (8.5%) | **<0.001** |
| Obesity | 346 (18.9%) | 435,650 (25.9%) | **<0.001** |
| Smoking | 617 (33.7%) | 444,060 (26.4%) | **<0.001** |
| Alcohol abuse | 55 (3.0%) | 45,415 (2.7%) | 0.20 |
| Pulmonary circulation disorders | 346 (18.9%) | 89,148 (5.3%) | **<0.001** |
| Peripheral vascular disorder | 340 (18.6%) | 80,738 (4.8%) | **<0.001** |
| Cardiac arrhythmias | 917 (50.1%) | 418,830 (24.9%) | **<0.001** |
| Congestive heart failure | 1,263 (69.0%) | 297,722 (17.7%) | **<0.001** |
| Chronic pulmonary disease | 372 (20.3%) | 373,414 (22.2%) | **<0.001** |
| Renal failure | 620 (33.9%) | 358,275 (21.3%) | **<0.001** |
| Liver disease | 183 (10.0%) | 92,512 (5.5%) | **<0.001** |
| AIDS/HIV | 9 (0.5%) | 3,365 (0.2%) | 0.41 |
| Rheumatoid arthritis/Collagen<br>vascular disease | 40 (2.2%) | 48,780 (2.9%) | 0.11 |
| Blood loss anemia | 20 (1.10%) | 6,728 (0.4%) | 0.08 |
| Deficiency anemia | 92 (5.0%) | 65,600 (3.9%) | **<0.001** |
| Solid tumor without<br>metastasis | 75 (4.1%) | 40,370 (2.4%) | **<0.001** |
| Metastatic cancer | 46 (2.5%) | 18,502 (1.1%) | **<0.001** |
| Lymphoma | 9 (0.5%) | 11,774 (0.7%) | 0.05 |

**Abbreviation:** IQR: Interquartile range

**Table 2:** Incidence of outcomes in overall COVID-19 cohort, COVID-19 with and without intracardiac thrombosis.

| **Outcomes** | **Overall cohort** | **COVID-19 without**<br>**intracardiac thrombosis** | **COVID-19 with**<br>**intracardiac thrombosis** | **p-value** |
| --- | --- | --- | --- | --- |
| In-hospital mortality | 13.2% (222,695/1,683,785) | 13.2% (222,385/1,682,045) | 16.9% (310/1,830) | **0.007** |
| Cardiovascular mortality | 3.5% (59,530/1,683,785) | 3.5% (59,370/1,682,045) | 8.7% (160/1,830) | **0.001** |
| Cardiac arrest | 2.6% (44,145/1,683,785) | 2.6% (44,055/1,682,045) | 4.9% (90/1,830) | **0.003** |
| Acute coronary syndrome | 4.4% (74,940/1,683,785) | 4.4% (74,670/1,682,045) | 14.8% (270/1,830) | **<0.001** |
| Heart failure | 16.1% (272,140/1,683,785) | 16.1% (270,955/1,682,045) | 64.8% (1,185/1,830) | **<0.001** |
| Stroke | 1.3% (22,030/1,683,785) | 1.2% (21,845/1,682,045) | 10.1% (185/1,830) | **<0.001** |
| Acute kidney injury | 28.3% (476,820/1,683,785) | 28.2% (475,955/1,682,045) | 47.3% (865/1,830) | **<0.001** |

### Factors associated with intracardiac thrombosis

Table 3 shows the multivariable analysis results for factors associated with intracardiac thrombosis. Female patients had a lower risk of intracardiac thrombosis (OR: 0.46, 95%CI: 0.36-0.58, p<0.001). Amongst racial groups, Native Americans had the highest risk (OR: 4.93, 95%CI: 2.32-10.47, p<0.001), followed by African Americans (OR: 2.68, 95%CI: 2.06-3.50, p<0.001). The clinical comorbidities most strongly associated with intracardiac thrombosis include a history of congestive heart failure (OR: 7.44, 95%CI: 5.37-10.32, p<0.001) and coagulopathy (OR: 1.70, 95%CI: 1.23-2.37, p=0.001). Compared to asymptomatic COVID-19 infection, symptomatic patients had a lower risk (OR: 0.62, 95%CI: 0.50-0.77, p<0.001), while those with severe symptoms had a higher risk (OR: 2.11, 95%CI: 1.32-3.37, p<0.001) of intracardiac thrombosis.

**Table 3:** Predictors of intracardiac thrombosis in COVID-19 patients.

| Predictors | OR (95%CI) | P-value |
| --- | --- | --- |
| **Demographic** |  |  |
| Age (per 10 years) | 0.99 (0.99-1.00) | 0.35 |
| Female | 0.46 (0.36-0.58) | **<0.001** |
| Race |  |  |
| Caucasian | Reference |  |
| African American | 2.68 (2.06-3.50) | **<0.001** |
| Hispanic | 2.28 (1.65-3.14) | **<0.001** |
| Asian or Pacific Islander | 2.54 (1.51-4.28) | **<0.001** |
| Native American | 4.93 (2.32-10.47) | **<0.001** |
| Other | 2.54 (1.51-4.28) | **<0.001** |
| Region |  |  |
| Northeast | Reference |  |
| Midwest | 1.53 (1.06-2.22) | **0.02** |
| South | 1.06 (0.74-1.52) | 0.75 |
| West | 1.10 (0.75-1.64) | 0.61 |
| Hospital location |  |  |
| Rural | Reference |  |
| Urban non-teaching | 1.33 (0.72-2.45) | 0.35 |
| Urban teaching | 2.13 (1.24-3.65) | **0.006** |
| Hospital size |  |  |
| Small | Reference |  |
| Medium | 1.32 (0.94-1.85) | 0.11 |
| Large | 1.57 (1.54-2.14) | **0.004** |
| **Presentation** |  |  |
| Asymptomatic | Reference |  |
| Symptomatic | 0.62 (0.50-0.77) | **<0.001** |
| Severely symptomatic | 2.11 (1.32-3.37) | **0.002** |
| **Comorbidities** |  |  |
| Hypertension | 0.92 (0.67-1.27) | 0.63 |
| Diabetes mellitus | 0.85 (0.67-1.07) | 0.17 |
| Hyperlipidemia | 1.05 (0.83-1.33) | 0.68 |
| Obesity | 0.81 (0.61-1.08) | 0.14 |
| Alcohol abuse | 0.34 (0.18-0.65) | **0.001** |
| Smoking | 0.81 (0.64-1.03) | 0.08 |
| Congestive heart failure | 7.66 (5.37-10.9) | **<0.001** |
| Cardiac arrhythmias | 1.58 (1.12-2.23) | **0.01** |
| Atrial fibrillation | 0.96 (0.68-1.34) | 0.80 |
| Valvular disease | 0.48 (0.33-0.70) | **<0.001** |
| Peripheral Vascular disease | 1.35 (0.99-1.83) | 0.05 |
| Pulmonary circulation disease | 1.93 (1.39-2.69) | **<0.001** |
| Chronic pulmonary disease | 0.58 (0.44-0.76) | **<0.001** |
| Prior cerebrovascular accident | 1.59 (1.12-2.12) | **0.001** |
| Renal failure | 0.74 (0.57-0.96) | **0.02** |
| Liver disease | 1.24 (0.85-1.81) | 0.26 |
| Coagulopathy | 1.67 (1.20-2.32) | **0.002** |
| Thrombophilia | 1.33 (1.20-1.47) | **<0.001** |
| AIDS/HIV | 1.01 (0.25-4.08) | 0.99 |
| Rheumatoid arthritis | 0.47 (0.21-1.05) | 0.06 |
| Blood loss anemia | 1.17 (0.43-3.16) | 0.75 |
| Deficiency anemia | 0.76 (0.48-1.22) | 0.26 |
| Solid tumor without metastasis | 0.72 (0.37-1.39) | 0.32 |
| Metastatic cancer | 0.82 (0.35-1.93) | 0.65 |
| Lymphoma | 0.37 (0.09-1.49) | 0.16 |

**Abbreviations:** OR: odds ratio; 95%CI: confidence interval

### Predictors of in-hospital mortality

Figure 1 illustrates the univariable and multivariable odds ratios of intracardiac thrombosis and in-hospital outcomes. Full multivariable analyses of results for in-hospital deaths are presented in Table 4. Intracardiac thrombosis was independently associated with a higher risk of in-hospital all-cause mortality (multivariable OR: 3.32, 95%CI: 2.42-4.54, p<0.001) and cardiovascular-related mortality (OR: 2.95, 95%CI: 1.96-4.44, p<0.001).Female patients had a lower risk of all-cause mortality (OR: 0.74, 95%CI: 0.73-0.75, p<0.001) and cardiovascular mortality (OR: 0.67, 95%CI: 0.66-0.69, p<0.001). Among all the racial and ethnic groups studied, Native Americans were associated with the highest risk of in-hospital mortality (OR: 4.93, 95% CI 2.32-10.47, p<0.001).Compared to asymptomatic patients, symptomatic patients (OR: 3.22, 95%CI: 3.09-3.37, p<0.001 and OR: 2.90 95%CI: 2.71-3.10, p<0.001) and those with severe symptoms (OR: 2.79, 95%CI: 2.71-2.88, p<0.001 and OR: 2.20, 95%CI: 2.11-2.30, p<0.001) had higher risk of all-cause and cardiovascular-related mortality, respectively. The most important predictors of all-cause mortality were coagulopathy (OR: 2.55, 95%CI: 2.50-2.60, p<0.001), liver disease (OR: 2.52, 95%CI: 2.47-2.57, p<0.001), and cardiac arrhythmias (OR: 2.09, 95%CI: 2.05-2.14, p<0.001). For mortality due to cardiovascular causes, the most important predictors were cardiac arrhythmias (OR: 5.12, 95%CI: 4.98-5.26, p<0.001), congestive heart failure (OR: 3.24 95%CI: 3.14-3.34, p<0.001), and liver disease (OR: 2.71, 95%CI: 2.64-2.79, p<0.001).

**Figure 1:**
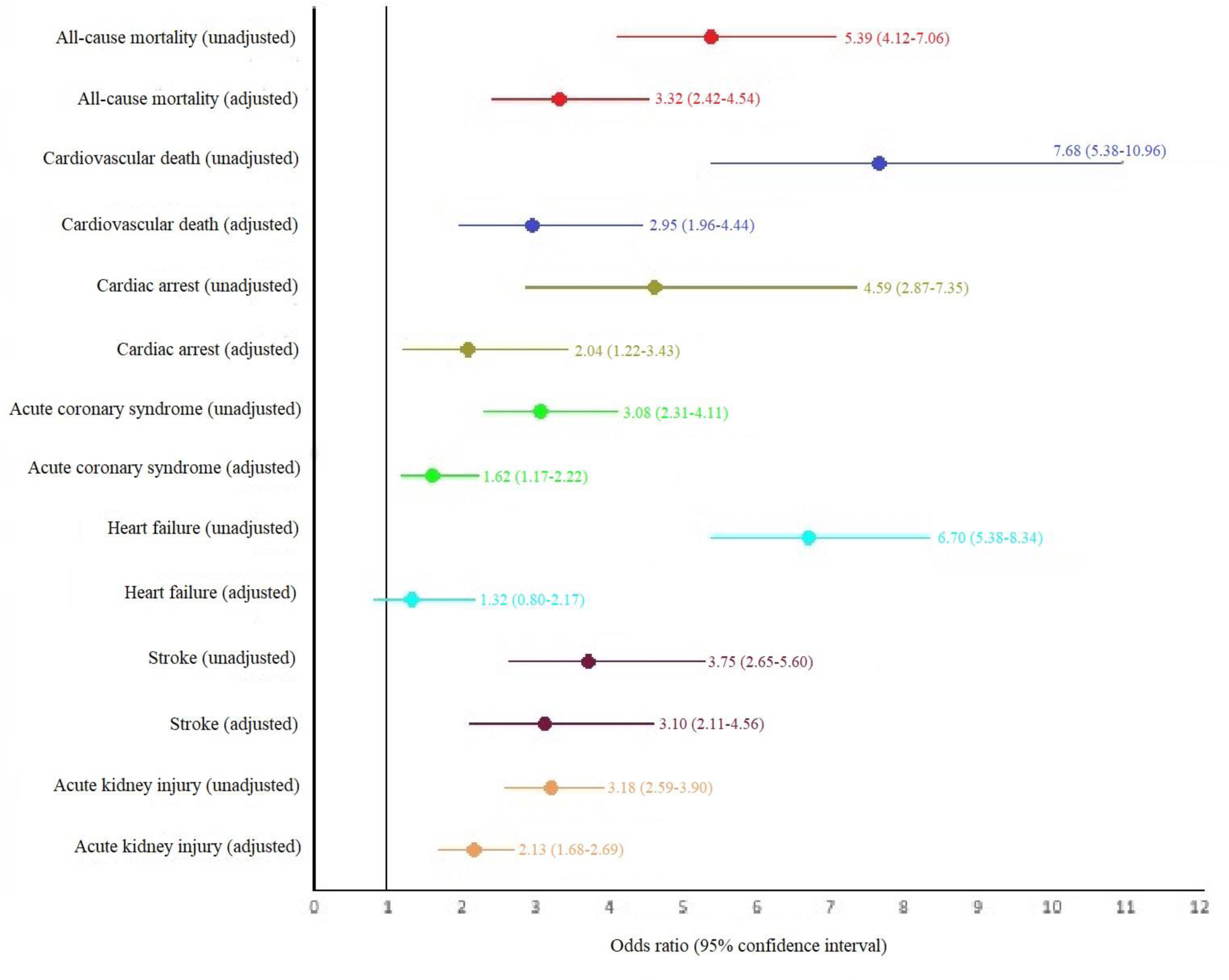
Forest plot depicting the unadjusted and adjusted odds ratio and 95% confidence interval for all the outcomes.

**Table 4:** Predictors of in-hospital mortality (all-cause and cardiovascular) in COVID-19 patients.

| Predictors | All-cause mortality |  | Cardiovascular mortality |  |
| --- | --- | --- | --- | --- |
|  | OR (95% CI) | P-value | OR (95% CI) | P-value |
| Intracardiac thrombus | 3.32 (2.42-4.54) | <0.001 | 2.95 (1.96-4.44) | **<0.001** |
| **Demographics** |  |  |  |  |
| Age (per 10 years) | 1.48 (1.34-1.50) | <0.001 | 1.21 (1.10-1.22) | **<0.001** |
| Female gender | 0.74 (0.73-0.75) | <0.001 | 0.67 (0.66-0.69) | **<0.001** |
| **Race** |  |  |  |  |
| Caucasian | Reference |  | Reference |  |
| African American | 1.22 (1.19-1.26) | <0.001 | 1.35 (1.31-1.40) | **<0.001** |
| Hispanic | 1.26 (1.23-1.30) | <0.001 | 1.32 (1.26-1.38) | **<0.001** |
| Asian or Pacific Islander | 1.20 (1.15-1.25) | <0.001 | 1.19 (1.12-1.26) | **<0.001** |
| Native American | 1.58 (1.44-1.75) | <0.001 | 1.41 (1.26-1.57) | **<0.001** |
| Other | 1.34 (1.27-1.40) | <0.001 | 1.39 (1.31-1.48) | **<0.001** |
| **Region** |  |  |  |  |
| Northeast | Reference |  | Reference |  |
| Midwest | 0.93 (0.88-0.98) | 0.006 | 1.06 (0.99-1.12) | 0.05 |
| South | 0.96 (0.92-1.01) | 0.140 | 1.19 (1.12-1.25) | **<0.001** |
| West | 0.92 (0.88-0.97) | 0.003 | 1.08 (1.01-1.14) | **0.01** |
| **Hospital size** |  |  |  |  |
| Small | Reference |  | Reference |  |
| Medium | 1.17 (1.12-1.22) | <0.001 | 1.16 (1.11-1.22) | **<0.001** |
| Large | 1.25 (1.20-1.31) | <0.001 | 1.17 (1.11-1.22) | **<0.001** |
| Hospital location |  |  |  |  |
| Rural | Reference |  | Reference |  |
| Urban non-teaching | 1.07 (1.01-1.12) | 0.014 | 1.24 (1.16-1.32) | **<0.001** |
| Urban teaching | 1.23 (1.17-1.29) | <0.001 | 1.36 (1.28-1.44) | **<0.001** |
| **Presentation** |  |  |  |  |
| Asymptomatic | Reference |  | Reference |  |
| Symptomatic | 3.22 (3.09-3.37) | <0.001 | 2.90 (2.71-3.10) | **<0.001** |
| Severely symptomatic | 2.79 (2.71-2.88) | <0.001 | 2.20 (2.11-2.30) | **<0.001** |
| **Comorbidities** |  |  |  |  |
| Hypertension | 0.70 (0.68-0.71) | <0.001 | 0.79 (0.77-0.81) | **<0.001** |
| Diabetes mellitus | 1.10 (1.09-1.12) | <0.001 | 1.17 (1.15-1.20) | **<0.001** |
| Hyperlipidemia | 0.69 (0.67-0.70) | <0.001 | 0.70 (0.69-0.72) | **<0.001** |
| Obesity | 0.91 (0.89-0.93) | <0.001 | 0.85 (0.83-0.88) | **<0.001** |
| Alcohol abuse | 0.71 (0.69-0.72) | <0.001 | 0.63 (0.60-0.65) | **<0.001** |
| Smoking | 0.60 (0.59-0.61) | <0.001 | 0.63 (0.61-0.64) | **<0.001** |
| Congestive heart failure | 1.38 (1.35-1.40) | <0.001 | 3.24 (3.14-3.34) | **<0.001** |
| Cardiac arrhythmias | 2.09-2.05-2.14) | <0.001 | 5.12 (4.98-5.26) | **<0.001** |
| Atrial fibrillation | 0.71 (0.69-0.72) | <0.001 | 0.43 (0.41-0.44) | **<0.001** |
| Valvular disease | 0.70 (0.69-0.71) | <0.001 | 0.80 (0.78-0.82) | **<0.001** |
| Peripheral Vascular disease | 1.03 (1.01-1.05) | 0.008 | 1.14 (1.10-1.17) | **<0.001** |
| Pulmonary circulation disease | 1.33 (1.30-1.35) | <0.001 | 1.25 (1.21-1.28) | **<0.001** |
| Chronic pulmonary disease | 1.13 (1.12-1.15) | <0.001 | 1.04 (1.01-1.06) | **0.001** |
| Prior cerebrovascular accident | 0.90 (0.88-0.91) | <0.001 | 0.82 (0.80-0.85) | **<0.001** |
| Renal failure | 1.14 (1.12-1.15) | <0.001 | 1.17 (1.14-1.19) | **<0.001** |
| Liver disease | 2.52 (2.47 (2.57) | <0.001 | 2.71 (2.64-2.79) | **<0.001** |
| Coagulopathy | 2.55 (2.50-2.60) | <0.001 | 2.24 (2.18-2.30) | **<0.001** |
| Thrombophilia | 0.97 (0.95-0.99) | 0.025 | 0.96 (0.94-0.98) | **<0.001** |
| AIDS/HIV | 1.41 (1.30-1.53) | <0.001 | 1.19 (1.05-1.35) | **0.007** |
| Rheumatoid arthritis | 0.92 (0.89-0.95) | <0.001 | 0.85 (0.81-0.90) | **<0.001** |
| Blood loss anemia | 0.68 (0.64-0.72) | <0.001 | 0.74 (0.67-0.81) | **<0.001** |
| Deficiency anemia | 0.67 (0.65-0.69) | <0.001 | 0.63 (0.60-0.65) | **<0.001** |
| Solid tumor without metastasis | 1.21 (1.18-1.24) | <0.001 | 1.03 (0.99-1.07) | 0.19 |
| Metastatic cancer | 2.01 (1.95-2.06) | <0.001 | 1.42 (1.35-1.49) | **<0.001** |
| Lymphoma | 1.16 (1.12-1.21) | <0.001 | 1.02 (0.95-1.09) | 0.65 |

**Abbreviations:** OR: odds ratio; 95%CI: confidence interval

### In-hospital complications and predictors

Multivariable analyses for in-hospital complications are presented in Table 5. Intracardiac thrombosis was associated with a higher risk of cardiac arrest (OR: 2.04, 95%CI: 1.22-3.43, p=0.006), ACS (OR: 1.62, 95%CI: 1.17-2.22, p=0.003), stroke (OR: 3.10, 95%CI: 2.11-4.56, p<0.001), and AKI (OR: 2.13 95%CI: 1.68-2.69, p<0.001) but was statistically not associated with heart failure (OR 1.32, 95% CI 0.80-2.17, p=0.27). Age per 10 years was associated with all in-hospital complications. Females had a lower risk of cardiac arrest, ACS, stroke and AKI but a higher risk of heart failure. The most important predictors for cardiac arrest were cardiac arrhythmia (OR: 4.89, 95%CI: 4.75-5.03, p<0.001) and liver disease (OR: 2.43, 95%CI: 2.36-2.59, p<0.001). For ACS, the most important predictors were congestive heart failure (OR: 2.83, 95%CI: 1.78-2.88, p<0.001) and cardiac arrhythmias (OR: 1.87 95%CI: 1.84-1.91, p<0.001). For heart failure, the most important predictors were pulmonary circulation disease (OR: 1.88, 95%CI: 1.82-1.94, p<0.001) and chronic pulmonary disease (OR: 1.68, 95%CI: 1.64-1.72, p<0.001).For stroke, the most important predictors were prior history of cerebrovascular accident (OR: 2.26, 95%CI: 2.21-2.30, p<0.001) and hypertension (OR: 2.22, 95%CI: 2.16-2.27, p<0.001). The most important predictors of AKI were renal failure (OR: 3.37, 95%CI: 3.32-3.42, p<0.001) and coagulopathy (OR: 1.69, 95%CI: 1.66-1.71, p<0.001).

**Table 5:** Predictors of in-hospital complications in COVID-19 patients.

| Predictors | Cardiac arrest |  | Acute coronary syndrome |  | Heart failure |  | Stroke |  | Acute kidney injury |  |
| --- | --- | --- | --- | --- | --- | --- | --- | --- | --- | --- |
|  | OR (95% CI) | P | OR (95% CI) | P | OR (95% CI) | P | OR (95% CI) | P | OR (95% CI) | P |
| Intracardiac thrombus | 2.04 (1.22-3.43) | **0.006** | 1.62 (1.17-2.22) | **0.003** | 1.32 (0.80-2.17) | 0.27 | 3.10 (2.11-4.56) | **<0.001** | 2.13 (1.68-2.69) | **<0.001** |
| **Demographics** |  |  |  |  |  |  |  |  |  |  |
| Female gender | 0.70 (0.68-0.71) | **<0.001** | 0.66 (0.66-0.67) | **<0.001** | 1.32 (1.29-1.34) | **<0.001** | 0.89 (0.88-0.90) | **<0.001** | 0.70 (0.69-0.70) | **<0.001** |
| Age (per 10 years) | 1.10 (1.10-1.21) | **<0.001** | 1.10 (1.10-1.21) | **<0.001** | 1.21 (1.10-1.22) | **<0.001** | 1.21 (1.10-1.22) | **<0.001** | 1.21 (1.10-1.22) | **<0.001** |
| **Race** |  |  |  |  |  |  |  |  |  |  |
| Caucasian | Reference |  | Reference |  | Reference |  | Reference |  | Reference |  |
| African American | 1.51 (1.46-1.57) | **<0.001** | 0.82 (0.80-0.85) | **<0.001** | 1.38 (1.33-1.44) | **<0.001** | 1.23 (1.20-1.26) | **<0.001** | 1.25 (1.23-1.26) | **<0.001** |
| Hispanic | 1.31 (1.25-1.38) | **<0.001** | 0.92 (0.87-0.97) | **0.001** | 1.18 (1.12-1.25) | **<0.001** | 0.86 (0.83-0.89) | **<0.001** | 0.88 (0.87-0.90) | **<0.001** |
| Asian or Pacific | 1.28 (1.20-1.37) | **<0.001** | 1.08 (1.03-1.13) | **0.001** | 0.98 (0.90-1.07) | 0.72 | 1.13 (1.08-1.19) | **<0.001** | 0.88 (0.86-0.90) | **<0.001** |
| Islander |  |  |  |  |  |  |  |  |  |  |
| Native American | 1.25 (1.11-1.41) | <**0.001** | 1.08 (0.97-1.20) | 0.17 | 1.12 (0.99-1.25) | 0.05 | 0.91 (0.82-1.01) | 0.079 | 0.97 (0.92-1.02) | 0.34 |
| Other | 1.54 (1.45-1.65) | <**0.001** | 1.13 (1.06-1.20) | <**0.001** | 1.16 (1.08-1.25) | <**0.001** | 1.06 (1.01-1.12) | **0.018** | 1.02 (1.01-1.06) | 0.03 |
| **Region** |  |  |  |  |  |  |  |  |  |  |
| Northeast | Reference |  | Reference |  | Reference |  | Reference |  | Reference |  |
| Midwest | 1.14 (1.07-1.22) | <**0.001** | 1.01 (0.94-1.10) | 0.73 | 0.99 (0.93-1.07) | 0.95 | 1.09 (1.04-1.16) | **0.001** | 0.99 (0.96-1.03) | 0.65 |
| South | 1.33 (1.25-1.42) | <**0.001** | 1.06 (0.99-1.15) | 0.10 | 1.08 (1.02-1.15) | **0.006** | 1.17 (1.11-1.24) | <**0.001** | 1.07 (1.04-1.11) | <**0.001** |
| West | 1.17 (1.09-1.26) | <**0.001** | 1.08 (1.01-1.17) | **0.04** | 1.24 (1.6-1.32) | <**0.001** | 1.17 (1.10 (1.23) | <**0.001** | 1.06 (1.03-1.10) | **0.001** |
| **Hospital location** |  |  |  |  |  |  |  |  |  |  |
| Rural | Reference |  | Reference |  | Reference |  | Reference |  | Reference |  |
| Urban non-teaching | 1.28 (1.19-1.38) | <**0.001** | 1.06 (0.98-1.14) | 0.15 | 0.72 (0.67-0.78) | <**0.001** | 1.15 (1.09-1.22) | <**0.001** | 1.13 (1.09-1.16) | <**0.001** |
| Urban teaching | 1.30 (1.21-1.39) | <**0.001** | 0.14 (1.06-1.22) | **0.001** | 0.78 (0.73-0.84) | <**0.001** | 1.44 (1.37-1.52) | <**0.001** | 1.17 (1.14-1.21) | <**0.001** |
| **Hospital size** |  |  |  |  |  |  |  |  |  |  |
| Small | Reference |  | Reference |  | Reference |  | Reference |  | Reference |  |
| Medium | 1.13 (1.07-1.19) | <**0.001** | 1.12 (1.05-1.19) | **0.001** | 0.94 (0.89-0.99) | **0.03** | 1.18 (1.13-1.23) | <**0.001** | 1.02 (0.99-1.05) | 0.10 |
| Large | 1.03 (0.98-1.08) | 0.31 | 1.13 (1.06-1.20) | <**0.001** | 0.95 (0.90-1.01) | 0.08 | 1.40 (1.34-1.46) | <**0.001** | 1.02 (0.99-1.05) | 0.09 |
| **Presentation** |  |  |  |  |  |  |  |  |  |  |
| Asymptomatic | Reference |  | Reference |  | Reference |  | Reference |  | Reference |  |
| Symptomatic | 2.57 (2.39-2.76) | <**0.001** | 1.27 (1.21-1.33) | <**0.001** | 0.97 (0.95-0.99) | **0.02** | 0.58 (0.54-0.62) | <**0.001** | 1.40 (1.37-1.43) | <**0.001** |
| Severely symptomatic | 1.86 (1.76-1.95) | <**0.001** | 1.48 (1.43-1.54) | <**0.001** | 1.28 (1.25-1.30) | <**0.001** | 0.59 (0.56-0.62) | <**0.001** | 4.06 (3.98-4.15) | <**0.001** |
| **Comorbidities** |  |  |  |  |  |  |  |  |  |  |
| Hypertension | 0.87 (0.85-0.90) | <**0.001** | 1.46 (1.44-1.49) | <**0.001** | 1.45 (1.41-1.50) | <**0.001** | 2.22 (2.16-2.27) | <**0.001** | 1.21 (1.20-1.23) | <**0.001** |
| Diabetes mellitus | 1.22 (1.19-1.24) | <**0.001** | 1.04 (1.03-1.05) | <**0.001** | 1.23 (1.20-1.26) | <**0.001** | 1.11 (1.10-1.13) | <**0.001** | 1.39 (1.38-1.40) | <**0.001** |
| Hyperlipidemia | 0.76 (0.75-0.78) | <**0.001** | 1.64 (1.61-1.67) | <**0.001** | 0.78 (0.76-0.79) | <**0.001** | 1.45 (1.43-1.48) | <**0.001** | 0.89 (0.88-0.90) | <**0.001** |
| Obesity | 1.00 (0.99-1.02) | 0.80 | 1.02 (1.01-1.04) | **0.009** | 1.48 (1.45-1.52) | <**0.001** | 0.89 (0.87-0.90) | <**0.001** | 1.18 (1.17-1.19) | <**0.001** |
| Alcohol abuse | 0.75 (0.72-0.78) | <**0.001** | 0.62 (0.61-0.64) | <**0.001** | 0.91 (0.88-0.95) | <**0.001** | 0.89 (0.87-0.92) | <**0.001** | 0.99 (98-1.01) | 0.77 |
| Smoking | 0.69 (0.67-0.71) | <**0.001** | 1.17 (1.15-1.19) | <**0.001** | 1.04 (1.02-1.06) | **0.001** | 1.00 (0.99-1.02) | 0.68 | 0.85 (0.84-0.85) | <**0.001** |
| Congestive heart | 1.60 (1.55-1.64) | <**0.001** | 2.83 (1.78-2.88) | <**0.001** | N/A |  | 0.70 (0.69-0.71) | <**0.001** | 1.28 (1.27-1.29) | <**0.001** |
| failure |  |  |  |  |  |  |  |  |  |  |
| Cardiac arrhythmias | 4.89 (4.75-5.03) | <**0.001** | 1.87 (1.84-1.91) | <**0.001** | 0.87 (0.84-0.89) | <**0.001** | 1.05 (1.02-1.07) | <**0.001** | 1.23 (1.22-1.25) | <**0.001** |
| Atrial fibrillation | 0.42 (0.41-0.43) | <**0.001** | 0.46 (0.46-0.47) | <**0.001** | 1.57 (1.53-1.62) | <**0.001** | 1.08 (1.06-1.11) | <**0.001** | 0.84 (0.83-0.85) | <**0.001** |
| Valvular disease | 0.77 (0.75-0.80) | <**0.001** | 1.31 (1.29-1.34) | <**0.001** | 1.44 (1.40-1.48) | <**0.001** | 1.13 (1.11-1.16) | <**0.001** | 0.88 (0.87-0.89) | <**0.001** |
| Peripheral Vascular disease | 0.97 (0.94-1.00) | 0.06 | 0.94 (0.92-0.96) | <**0.001** | 1.00 (0.98-1.03) | 0.842 | 0.97 (0.95-0.99) | **0.01** | 0.97 (0.96-0.98) | <**0.001** |
| Pulmonary circulation disease | 1.43 (1.38-1.48) | <**0.001** | 1.07 (1.04-1.09) | <**0.001** | 1.88 (1.82-1.94) | <**0.001** | 0.82 (0.80-0.85) | <**0.001** | 1.13 (1.12-1.14) | <**0.001** |
| Chronic pulmonary disease | 1.02 (1.00-1.05) | 0.05 | 0.77 (0.76-0.79) | <**0.001** | 1.68 (1.64-1.72) | <**0.001** | 0.58 (0.57-0.59) | <**0.001** | 0.92 (0.92-0.93) | <**0.001** |
| Prior cerebrovascular accident | 0.83 (0.80-0.86) | <**0.001** | 0.81 (0.80-0.82) | <**0.001** | 1.08 (1.05-1.11) | <**0.001** | 2.26 (2.21-2.30) | <**0.001** | 0.91 (0.90-0.92) | <**0.001** |
| Renal failure | 1.09 (1.06-1.11) | <**0.001** | 0.92 (0.90-0.93) | <0.001 | 1.53 (1.50-1.57) | <**0.001** | 0.64 (0.63-0.65) | <**0.001** | 3.37 (3.32-3.42) | <**0.001** |
| Liver disease | 2.43 (1.36-2.59) | <**0.001** | 1.07 (0.05-1.09) | <0.001 | 1.19 (1.15-1.23) | <**0.001** | 0.60 (0.58-0.62) | <**0.001** | 1.69 (1.66-1.71) | <**0.001** |
| Coagulopathy | 2.02 (1.97-2.08) | <**0.001** | 1.23 (1.20-1.25) | <0.001 | 0.94 (0.91-0.97) | <**0.001** | 0.89 (0.86-0.91) | <**0.001** | 1.80 (1.78-1.83) | <**0.001** |
| Thrombophilia | 0.95 (0.93-0.97) | <**0.001** | 0.99 (0.99-1.00) | 0.638 | 0.99 (0.98-1.01) | 0.49 | 1.23 (1.21-1.25) | <**0.001** | 0.95 (0.95-0.96) | <**0.001** |
| AIDS/HIV | 1.05 (0.92-1.20) | 0.49 | 0.91 (0.84-0.99) | 0.036 | 1.10 (0.95-1.27) | 0.22 | 1.02 (0.91-1.14) | 0.71 | 1.44 (1.38-1.50) | <**0.001** |
| Rheumatoid arthritis | 0.84 (0.79-0.89) | <**0.001** | 0.92 (0.90-0.95) | <0.001 | 1.03 (0.99-1.08) | 0.18 | 0.81 (0.78-0.84) | <**0.001** | 1.05 (1.03-1.06) | <**0.001** |
| Blood loss anemia | 0.77 (0.70-0.85) | <**0.001** | 0.74 (0.70-0.78) | <0.001 | 1.14 (1.04-1.25) | <**0.001** | 0.51 (0.47-0.55) | <**0.001** | 1.05 (1.02-1.08) | <**0.001** |
| Deficiency anemia | 0.73 (0.70-0.77) | <**0.001** | 0.81 (0.79-0.83) | <0.001 | 1.38 (1.33-1.43) | <**0.001** | 0.65 (0.63-0.67) | <**0.001** | 1.33 (1.31-1.35) | <**0.001** |
| Solid tumor without metastasis | 0.97 (0.92-1.01) | 0.13 | 0.70 (0.68-0.72) | <0.001 | 0.88 (0.84-0.92) | <**0.001** | 0.68 (0.66-0.70) | <**0.001** | 1.09 (1.07-1.10) | <**0.001** |
| Metastatic cancer | 1.13 (1.07-1.20) | <**0.001** | 0.78 (0.75-0.81) | <0.001 | 0.88 (0.83-0.93) | <**0.001** | 0.95 (0.92-0.99) | 0.22 | 1.22 (1.20-1.24) | <**0.001** |
| Lymphoma | 0.80 (0.74-0.88) | <**0.001** | 0.63 (0.60-0.66) | <0.001 | 0.93 (0.87-0.99) | **0.04** | 0.56 (0.52-0.60) | <**0.001** | 1.25 (1.22-1.28) | <**0.001** |

**Abbreviations:** OR: Odds Ratio; 95%CI: Confidence interval

## DISCUSSION

Our study studied the incidence of intracardiac thrombosis in patients with COVID-19 and its association with various comorbidities, end-organ damage, and patient mortality. The incidence of intracardiac thrombosis in patients with COVID-19 was found to be 0.001%. Key predictors of intracardiac thrombosis were identified. Intracardiac thrombosis was independently associated with a higher risk of in-hospital death (all-cause and cardiovascular) and most in-hospital complications (cardiac arrest, ACS, stroke, and AKI).

### Predictors of Intracardiac thrombosis in COVID-19

Apart from primarily infecting the respiratory tract, COVID-19 is widely recognized for its multisystemic involvement and a multitude of potentially devastating complications.^10^ Nearly 36% of patients with COVID-19 have some cardiovascular involvement, which can include myocardial infarction, cardiomyopathy, arrhythmias, and myocarditis, and these patients are found to have worse clinical outcomes.^10–12^ Dweck et al. found that 55% of the patients with COVID-19 have some abnormalities on echocardiography irrespective of comorbidities and symptoms.^13^ Abnormally elevated troponin levels have been found in 27% to 71% of patients with COVID-19, with nearly 40% having worse outcomes.^14, 15^ Many previous studies have also shown that COVID-19 infection causes a hypercoagulable state that can lead to venous and arterial thromboembolism.^16–18^

Previously, many patients with COVID-19 have been documented to develop intracardiac thrombosis.^4–8^ Intracardiac thrombosis is usually seen in patients with endocardial disease, prosthetic valves, intracardiac devices, contraction anomalies, or thrombophilia.^19^ However, occurrence of intracardiac thrombosis has also been documented in normally contracting heart in patients with COVID-19.^7, 19^ Cases have been reported with biatrial thrombosis, biventricular thrombosis, and valvular thrombosis without any signs of infective endocarditis or coagulopathy.^19–21^ These thrombotic complications have occurred despite the early pharmacological thromboprophylaxis administration in hospitalized COVID-19 patients. Previous observational studies have shown the incidence of intracardiac thrombosis in 17 out of 1,010 patients (0.017%) and 5 out of 630 patients (0.008%) with COVID-19.^6, 22, 23^ Yuan et al performed echocardiography on all hospitalized patients with COVID-19 and found intracardiac thrombus in 2 out of 434 patients (0.0046%).^24^

Our large-scale nationwide study showed the incidence of intracardiac thrombosis in COVID-19 inpatients was 0.001%, which although rare may have important clinical consequences. The presence of multiple factors associated with intracardiac thrombosis, including male sex, non-Caucasian ethnicities especially Native Americans, heart failure, prior stroke and coagulopathy, should alert clinicians to a higher suspicion for intracardiac thrombosis if there are concerning clinical symptoms or deterioration in COVID-19 inpatients. Women having lower risk than men for intracardiac thrombosis is interesting, and this might be explained by the gender differences in the expression of ACE-2 receptors due to higher levels of androgens in men. These surface receptors line the endothelial cells and cells in other organ systems and are a site for viral entry.^25^ Furthermore, gender differences in innate immunity may also be involved.^26^

### Adverse outcomes of intracardiac thrombosis in COVID-19

In our study, intracardiac thrombosis was found to be associated with higher risk of all-cause and cardiovascular mortality, cardiac arrest, ACS, stroke, and AKI, highlighting the clinical significance of this finding. The common and most plausible mechanism is of course an embolic event to the coronary, carotid, and renal vessels respectively, resulting in a life-threatening complication. Severely symptomatic COVID-19 was also associated with heart failure in our analyses, however given that history of heart failure is an established risk factor of intracardiac thrombosis (especially in the left ventricle), it is more likely that heart failure precedes intracardiac thrombosis. A recent study showed worse clinical outcomes in ST-elevation myocardial infarction (STEMI) patients with COVID-19 with higher overall thrombus burden and higher incidence of cardiac arrest, multivessel thrombosis, stent thrombosis, and in-hospital mortality compared to STEMI patients without COVID-19.^27^

Interestingly, men had higher risk of all in-hospital complications except heart failure. Despite having higher risk of ACS, stroke, and AKI than some of the other races, our study found that Caucasian patient population had the lowest risk of mortality amongst all the races. African American patients had the highest risk for heart failure, stroke, and AKI, while patients belong to the group of “other” races had the highest risk of cardiac arrest and ACS. These disparities might be explained due to the genetic differences, differences in general health status, health-associated social factors, and inequity in access to healthcare services.^28, 29^

### Clinical implications

Intracardiac thrombosis is more frequently found in left-sided cardiac chambers compared to the right side. Lee et al reported 130 events of intracardiac thrombosis after screening 86,374 patients with echocardiography out of which 47.4% were located in left ventricle (LV), 37.7% in left atrium (LA), 11.5% in the right atrium (RA) and 3.1% in the right ventricle (RV).^30^ The main causes of LV thrombus include ST-elevation myocardial infarction (STEMI) (incidence: 2.9%-15%) and non-ischemic cardiomyopathies (incidence: 2-36%), while LA thrombus is most frequently seen in patients with atrial fibrillation (AF) (incidence: 5%-27% without anticoagulation), with 90% occurring in the left atrial appendage (LAA).^31–, 34^ Deep venous thrombosis (DVT) is the most common cause of thrombosis in right cardiac chambers.^35^ Abnormal elevation of certain lab values has been shown to predict a higher risk of intracardiac thrombosis in COVID-19 patients, including ALT, D-dimer, proBNP, and HS troponin T.^23^

Transthoracic echocardiography (TTE) is usually the first imaging technique for bedside assessment of intracardiac thrombus.^36^ However, TTE can be difficult in patients with small intercostals spaces, large body habitus, lung diseases leading to poor acoustic windows. Transesophageal echocardiography (TEE) can be used in such patients due to its better visualization of the left atrium (which is directly adjacent to the esophagus) and thus higher sensitivity for detecting LAA thrombus.^37^ This can be supplemented by the use of echocardiographic contrast (ECA) and harmonic imaging to further improve the sensitivity and specificity. Studies have shown that the use of ECA can improve the sensitivity from 33% to 100% and specificity from 82% to 92% in LVT detection.^38, 39^ Cardiac MR with gadolinium contrast is another imaging technique that can be used for detection of intracardiac thrombus along with other structural cardiac anomalies. A study showed that cardiac MR had the highest sensitivity (88%) and specificity (99%) compared to TEE (40% and 96%) and TTE (23% and 96%) for detection of LVT.^37, 40^ However, the high cost and less availability limits the broad use of cardiac MR. Contrast-enhanced CT is another technique that can be used with 65 Hounsfield units as a threshold for intracardiac thrombus.^41^ The limitations include radiation exposure and use of intravenous contrast.

It is also important to ensure appropriate transmission prevention in COVID-19 patients while performing these studies. European Society of Cardiology (ESC) and the American Society of Echocardiography (ASE) guidelines do not recommend echocardiography being routinely performed in patients with COVID-19.^42, 43^ Screening should be clinically driven based on symptoms. Anticoagulation is the mainstay of treatment. Vitamin-K antagonists (VKAs) have been conventionally used for a period 3-6 months.^44^ Recently, direct oral anticoagulants (DOACs) have also been used to treat intracardiac thrombus. Despite the lack of randomized controlled trials (RCTs) data, DOACs have several advantages over VKAs: rapid onset of action, short half-lives permitting fixed dose regimens, oral route of administration, and lack of requirement of lab monitoring.^45^ Recent studies have shown thrombus resolution rates ranging from 83% to 90% with lower risks of bleeding and variable risk of embolization.^45–47^ The American Heart Association recommends the use of low molecular heparin, dabigatran, apixaban, and rivaroxaban in cases of warfarin intolerance.^44^ The duration of therapy depends on the etiology and risk of thrombosis. Patients with reversible causes such as infection, resolved cardiomyopathy, or atrial fibrillation with low CHA2DS2-VASC scores require shorter duration compared to those with persistent risk factors. In hemodynamically unstable patients, surgical thrombectomy may be used on a case-to-case basis.^44^

### Limitations

Our study has some limitations. This is a retrospective observational study with intrinsic biases but represents amongst the largest COVID-19 experience to date evaluating intracardiac thrombosis as part of the NIS database. The true prevalence of intracardiac thrombosis is likely higher than reported, as most patients would not have undergone all the tests for detecting thrombosis, and the method of evaluation may differ in utility and accuracy in different healthcare practices. The type of intracardiac thrombosis, whether left atrial appendage, left or right atrial or ventricular cannot be distinguished via the diagnostic coding of the NIH database for subgroup analyses. The time course of the development of intracardiac thrombosis to the COVID-19 infection is unknown, and may have been causally related, incidental, or even pre-existing. Studies have shown that intracardiac thrombus may be found from 5 to 25 days and up to 80 days in cases of delayed thrombosis.^48, 49^ Patients may develop intracardiac thrombosis after being treated for acute COVID-19 illness and discharged from the hospital. Our study lacks data on those patients since NIS does not track patients’ post-discharge. Lastly, we lack the data on laboratory tests results, imaging modalities and treatments used in the NIS dataset to allow for analyses.

## Conclusion

In conclusion, incidence of intracardiac thrombosis was 0.001% in COVID-19 in this large representative US inpatient sample. Intracardiac thrombosis was associated with a higher risk of mortality (all-cause and cardiovascular) and some in-hospital complications (cardiac arrest, ACS, stroke, and AKI). Key factors associated with intracardiac thrombosis include male sex, non-Caucasian ethnicities especially Native Americas, heart failure, prior stroke and coagulopathy, and the presence of several of these factors should alert clinicians to a higher suspicion for intracardiac thrombosis if there are concerning clinical symptoms or deterioration in COVID-19 inpatients.

## Data Availability

Data related to the manuscript is with the corresponding author and is available on request.

## Notes

### Competing Interest Statement

The authors have declared no competing interest.

### Funding Statement

No funding received in any form.

### Author Declarations

Publicly available and anonymous data from the National Inpatient Sample database were used for our study, which precludes the requirement of Institutional Review Board approval and informed consent.

